# Immune recovery in tumor microenvironment of *TP53*-mutated AML following venetoclax combination therapy

**DOI:** 10.1101/2025.05.12.25327414

**Authors:** Kohei Yamada, Jun Ando, Yoshiki Furukawa, Midori Ishii, Shintaro Kinoshita, Kota Tachibana, Yoko Azusawa, Yoko Edahiro, Yutaka Tsukune, Hajime Yasuda, Tomoiku Takaku, Yasuharu Hamano, Makoto Sasaki, Masanori Nojima, Miki Ando

**Author notes:** Correspondence: Miki Ando, M.D., Ph.D., M.A., Department of Hematology, Juntendo University School of Medicine, 2-1-1 Hongo, Bunkyo-ku, Tokyo 113-8421, Japan.

## Abstract

**Highlights:**

1. Single-cell RNA sequencing of bone marrow from TP53-mutated AML patients showed a decrease in cells expressing anti-apoptotic genes like BCL2 and MCL1 after venetoclax and azacitidine treatment.
2. Immune cells increased both in number and in gene expression levels associated with cytotoxicity post venetoclax combination therapy, verifying immune recovery.
3. Residual AML cells expressed CD47 and CLL1, suggesting a role for ancillary treatment targeting antigens expressed on residual *TP53*-mutated AML cells.

The BCL-2 inhibitor venetoclax combined with the hypomethylating agent azacitidine or with low-dose cytarabine significantly improves response rates and overall survival (OS) for newly diagnosed unfit and relapsed / refractory (R/R) acute myeloid leukemia (AML) patients. We retrospectively analyzed our experience with venetoclax combination therapy in 41 unfit AML patients (23 untreated, 18 R/R). Overall response rates were 78.3% for untreated patients and 61.1% for R/R patients. *TP53* alterations were observed in 13 patients (31.7%) and were identified as an independent predictor of poor outcome (p=0.0008). We further conducted single-cell RNA sequencing in bone marrow, sampled before and after venetoclax and azacitidine treatment, of three *TP53*-mutated AML patients who achieved complete remission (CR) or CR with incomplete hematologic recovery. After treatment, numbers of cells expressing anti-apoptotic genes such as *BCL2* and *MCL1* decreased. CD4 T cells, cytotoxic CD8 T cells, and NK cells significantly increased both in number and in levels of gene expression associated with cytotoxicity post-treatment, confirming immune recovery in the tumor microenvironment. Residual AML cells expressed *CD47* and *CLEC12A* (CLL1). These results indicate that venetoclax combination therapy of AML is promising in real-world clinical practice and suggest a role for ancillary treatment targeting antigens expressed on residual AML cells as a therapeutic strategy in *TP53*-mutated AML.

## Introduction

Acute myeloid leukemia (AML) is primarily a disease of elderly patients, with a median age at diagnosis of 68 years ^1,2^. Unfortunately, standard treatment involves intensive chemotherapy and allogeneic hematopoietic stem cell transplantation. Thus it is for many patients not an option because of advanced age, comorbidities, performance status, and increased likelihood of treatment resistance, resulting in poor prognosis ^3–6^. Such patients, being unfit for standard treatment, have received less intensive therapy, such as the hypomethylating agents azacitidine and decitabine, or low-dose arabinosylcytosine / cytarabine (LDAC)^7,8^. However, use of the highly selective oral B-cell leukemia/lymphoma-2 (BCL-2) inhibitor venetoclax combined with these hypomethylating agents recently has yielded remarkable results that have substantially reshaped AML management ^9,10^. Phase 3 studies in newly diagnosed elderly/unfit patients have demonstrated efficacy of venetoclax against AML in combination with hypomethylating agents ^11^ or with LDAC ^12^, permitting active treatment of relapsed/refractory (R/R) AML patients as well, albeit with highly variable results not so good as those for newly diagnosed unfit AML patients ^13–20^. Furthermore, in R/R patients such treatment often is more hematologically toxic, with febrile neutropenia (FN) predominating. Resultant long-term hospitalization impairs quality of life ^21,22^. To understand better the clinical response and survival rates in a real-world population of untreated AML and R/R AML patients given venetoclax combination therapy, we retrospectively conducted a single-center analysis at our institution to elucidate clinical and molecular characteristics, with associated clinical outcomes. In addition, we sought clinical and genetic biomarkers associated with benefits of venetoclax combination therapy for untreated and R/R patients using next generation sequencing (NGS) and performed subgroup analyses. We identified *TP53* alterations (*TP53* mutation and loss of chromosome 17) as independent predictors of poor outcome.

We further performed single-cell RNA sequencing (scRNA-seq) of pre- and post-treatment bone marrow samples from three *TP53*-mutated AML patients who achieved complete remission (CR) or CR with incomplete hematologic recovery (CRi) to understand the tumor microenvironment in these instances of favorable outcome.

## Methods

### Ethics statement

The Research Ethics Committee, Faculty of Medicine, Juntendo University School of Medicine approved “Genetic Mutation Analysis of Hematological Diseases” for this study on 1 October 2012 (approval number: M12-0866) and “Retrospective Study on Venetoclax Combination Therapy for Acute Myeloid Leukemia” for this project on 21 April 2022 (approval number: E22-0051). This study was conducted in accordance with the Declaration of Helsinki. All participants provided written informed consent.

### Patient characteristics

After approval of the Ethics Committee at Juntendo University School of Medicine, 41 patients with AML (23 previously untreated and 18 R/R) who at study onset were venetoclax-naïve and who had received venetoclax and azacitidine or venetoclax and LDAC during September 2019 to April 2023 were retrospectively recruited. Median follow-up was 258 days for untreated patients and 275 days for R/R patients. Three patients who had previously received azacitidine were treated with venetoclax and LDAC. Cytogenetic, molecular, and immunophenotypic data were collected at initiation of venetoclax therapy and at relapse. AML type was assigned by WHO 2016 criteria and 2017 European LeukemiaNet (ELN) risk stratification was used to evaluate risk groups ^23^. “Complex karyotype” was defined as 3 or more chromosomal abnormalities in the absence of one of the WHO-designated recurring translocations or inversions ^23^.

### Treatment characteristics

Patients were given a standardized-protocol dose of venetoclax following practice guidelines ^24^. All patients were hospitalized during the first cycle for tumor lysis syndrome prophylaxis. Granulocyte colony stimulating factor (G-CSF) was administered according to institutional practice based on VIALE studies ^11,12^. Among our patients, 9 patients (4 of 23 untreated and 5 of 18 R/R, respectively 17.3% and 27.8%) underwent subsequent hematopoietic stem cell transplantation (HSCT).

### Molecular characteristics

DNA and RNA extraction for molecular analyses of bone marrow used Nucleospin TriPrep (Takara Bio, Shiga, Japan). DNA concentrations, quantified using a Qubit 3 fluorometer (Thermo Fisher Scientific, Waltham, MA), were adjusted to 50ng/µl. Tumor profiling to detect sequence alterations and gene fusions was conducted by Oncomine^TM^ Childhood Cancer Research Assay (Thermo Fisher Scientific) following. The assay addresses specific mutations, gene amplifications, and fusions in a panel of 203 genes representing multiple classes.

Sequencing was performed using the Ion Chef^TM^ Instrument (Thermo Fisher Scientific). *FLT3*/ITD mutation was detected using Takara PCR *FLT3*/ITD Mutation Detection Kit (Takara Bio). All equipment was used following manufacturer instructions. *FLT3*/ITD allele frequency was calculated as the area under the curve of a mutant allele as a percentage of mutant and wild-type alleles. With more than one mutant allele, the values were aggregated. Mutant length was calculated by subtracting number of bases with wild-type *FLT3* from number of bases containing mutant *FLT3*.

### Response criteria and survival

Overall survival (OS) was defined as interval from diagnosis to last follow-up or to death from any cause. Overall response rate (ORR) was defined as CR or CRi. Response was assessed by 2017 ELN response criteria ^23^.

### Safety analyses

Hematologic and non-hematologic toxicity was evaluated by Common Terminology Criteria and Adverse Events classification (CTCAE) version 5.0.

### Single-cell RNA sequencing

RNA was extracted using TRIzol Reagent (Invitrogen). ScRNA-seq was performed using the 10x Chromium system (10x Genomics, Pleasanton, CA). In brief, approximately 2.0 x 10^6^ cells, viability >85%, were suspended in 50μl of chilled phosphate-buffered saline + 0.04% bovine serum albumin (Thermo Fisher Scientific) and processed to yield scRNA-seq libraries using 10X Chromium Next Gem Single Cell 3’ Reagent Kits v 3.1 (Dual Index) (10x Genomics). A Bio-Rad T100 thermal cycler (Bio-Rad Laboratories, Hercules, CA) was used for PCR. After the libraries were qualified using Bioanalyzer 2100 (Agilent, Tokyo, Japan), samples were subjected to sequencing (Macrogen, Seoul, South Korea) on the HiseqX (Illumina, San Diego, CA) platform at a depth of 2,000,000 pair reads/sample. Raw reads (fastq files) were re-aligned using Cell Ranger (v7.0.0) (10x Genomics). Normalization and downstream analysis of RNA data were performed using the Seurat R package (version 4.3.0)^25^, which enables the integrated processing of multi-modal single cell datasets. Clusters were manually annotated using published gene expression profiles ^26,27^ and further annotated using the FindAllMarkers functions. Differential gene expression analysis between clusters or cell populations in samples used the FindMarkers function (Wilcoxon rank-sum test).

### Statistics

OS and PFS were evaluated by the Kaplan-Meier method with differences between groups compared by log-rank test; *p* values of <0.05 were considered statistically significant. Statistical analyses were performed using GraphPad Prism 9 (GraphPad, San Diego, CA).

Multivariate analysis for OS was conducted by Cox regression. Backward elimination method (excluded when P>0.1) was applied due to the limited number of events. The variables included in the Cox regression model as potential prognostic factors were age, sex, performance status, blast count (>20%), prior treatment, HSCT, CR, complex karyotypes, *TP53* alterations and ELN risk stratification. Statistical analyses were performed using GraphPad Prism 9 and SPSS 25 (IBM, Armonk, NY).

## Results

### Patient characteristics

Forty-one adult AML patients (median age 71 years, range 40-87; 23 untreated, 18 R/R) received venetoclax combination therapy (Figure 1A and Table 1). AML was *de novo* in 8 (19.5%; untreated: 1 patient, R/R: 7 patients), secondary in 33 (80.5%; untreated: 22 patients, R/R: 11 patients), and therapy-related in 5 (12.2%; untreated: 4 patients, R/R: 1 patient). Twenty-seven (65.9%; untreated: 18 patients, R/R: 9 patients) had before been diagnosed with myelodysplastic syndrome (MDS) / AML with myelodysplasia-related changes (AML-MRC). Risk stratification according to 2017 ELN criteria ^23^ classed disease in 5 patients as favorable (12.2%), in 9 as intermediate (22.0%), and in 26 as adverse (63.4%) patients (Table 1).

**Figure 1.**
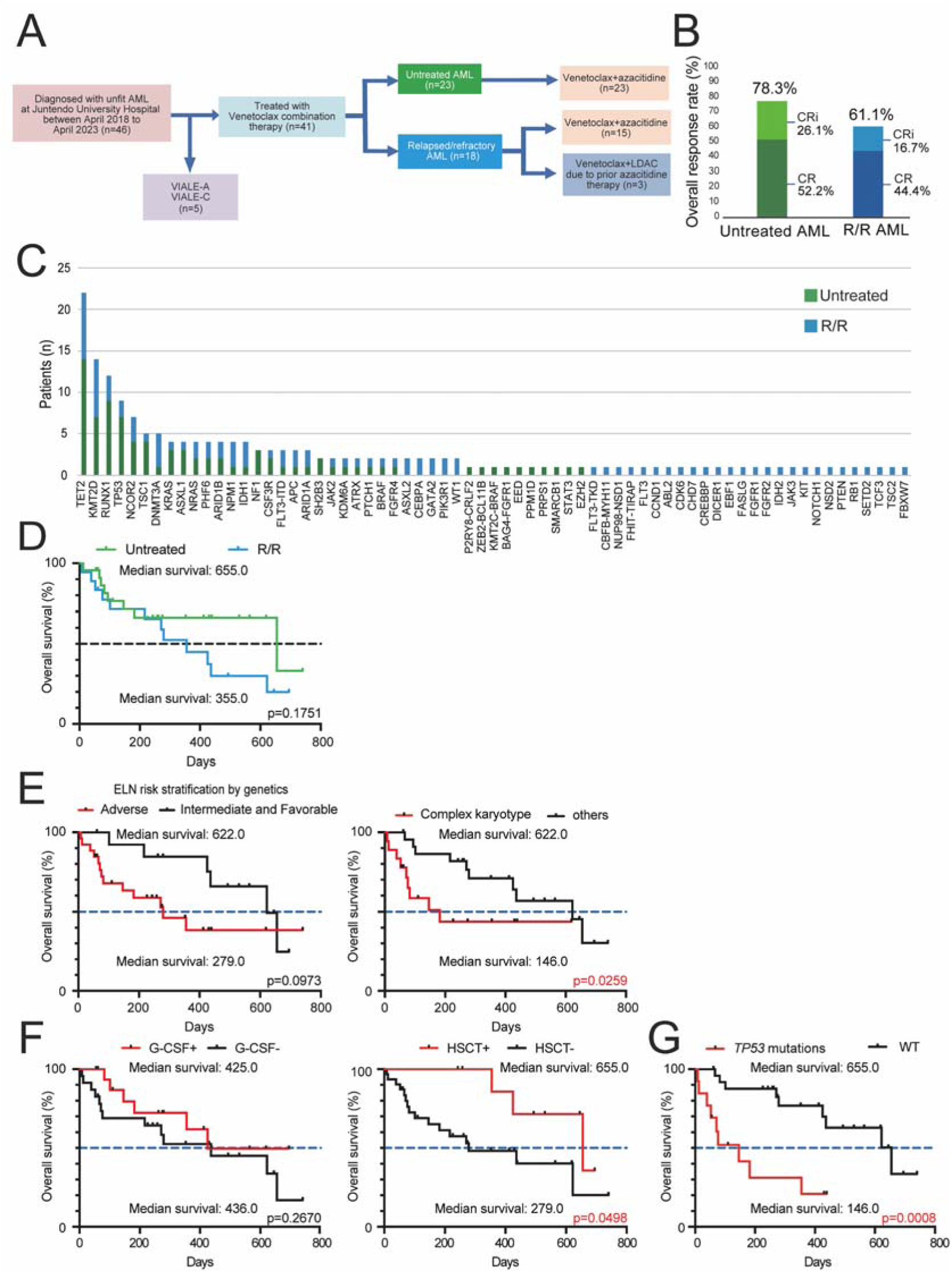
Overall response rates, Kaplan-Meier analyses showing overall survival and detected mutations. (A) Study design. (B) Overall response rates of untreated AML and R/R AML patients. Untreated AML patients; CR, dark green; CRi, light green. R/R AML patients; CR, dark blue; CRi, light blue. (C) Bar graph, numbers of patients with detected mutations. Untreated patients, green; R/R patients, blue. (D) Kaplan-Meier analysis showing overall survival. Overall survival of untreated and R/R AML patients. (E) Subgroup analyses of overall survival classed by ELN risk stratification and presence of complex karyotype. (F) Subgroup analyses of overall survival presence of G-CSF treatment and HSCT. (G) Subgroup analysis of overall survival presence of *TP53* alterations.

**Table 1.** Patient characteristics.

| Characteristic |  | Untreated AML,<br>n=23 | Relapsed / Refractory AML,<br>n=18 |
| --- | --- | --- | --- |
| **Sex, n (%)** | Female | 9 (39) | 10 (56) |
|  | Male | 14 (61) | 8 (44) |
| **Age, n (%)** | Median (range) | 74 (87-52) | 68 (84-40) |
|  | <60 | 2 (9) | 6 (33) |
|  | 60-75 | 12 (52) | 9 (50) |
|  | >75 | 9 (39) | 3 (17) |
| **ECOG PS, n (%)** | 0-1 | 19 (83) | 18 (100) |
|  | ≥2 | 4 (17) | 0 (0) |
| **Type of AML, n (%)** | De novo | 1 (4) | 7 (39) |
|  | Secondary | 22 (96) | 11 (61) |
|  | Previous MDS/AML-MRC | 18 (78) | 9 (50) |
|  | Therapy related | 4 (17) | 1 (6) |
| **ELN risk, n (%)** | Favorable | 1 (4) | 4 (22) |
|  | Intermediate | 4 (17) | 5 (28) |
|  | Adverse | 17 (74) | 9 (50) |
| **Treatment, n (%)** | Venetoclax + Azacitidine | 23 (100) | 15 (83) |
|  | Venetoclax + LDAC | 0 (0) | 3 (17) |

### Treatment characteristics

Venetoclax and azacitidine were administered to 38 patients (91.6%), 23 untreated and 15 R/R, respectively 100% and 83%. The remaining 3 R/R patients (17%) were given venetoclax and LDAC because they had received azacitidine before (Figure 1A and Table 1). Within the R/R patient group, prior therapy included idarubicin and cytarabine (66.7%), azacitidine monotherapy (27.8%), low-dose cytarabine, aclarubicin hydrochloride, and G-CSF (22.2%), gemutuzumab ozogamicin (11.1%), pevonedistat (5.6%), high-dose cytarabine (5.6%), HSCT (16.7%), and lenalidomide (5.6%). All patients were treated according to VIALE study protocols^11,12^ with venetoclax administered orally using a 28-day cycle for most patients (63.4%). Thirty-five (85.4%) received 400 mg venetoclax; one (2.4%) was given 50 mg since he was receiving azole antifungal prophylaxis; and 3 (7.3%) received 200 mg or 300 mg venetoclax because of prolonged bone marrow suppression. The remaining 3 patients (7.3 %) who were treated with LDAC received 600mg venetoclax according to the VIALE-C study protocol. The median length of venetoclax administration was 28 days (range 7-28). The average number of courses of venetoclax treatment was 3.93 (range 1-14); 18 patients (43.9%) received 3 or more cycles of therapy.

### Response to treatment

Overall response rate (ORR) was 78.3% for untreated patients, with 52.2% CR and 26.1% CRi. ORR was 61.1% for R/R patients, with 44.4% CR and 16.7% CRi (Figure 1B).

### Toxicity assessment

Grade 3 and 4 hematological adverse effects are listed in Table 2. Neutropenia was seen in all patients. FN occurred in 19 untreated patients (82.6%) and 7 R/R patients (38.9%), and was treated with antibiotics; no patients died directly because of FN. Although anti-fungal prophylaxis was given to only 2 patients (4.8%), both R/R, had invasive fungal infection. One suffered from candidemia and the other from mucormycosis (Table 2).

**Table 2.** Notable hematological or infectious adverse events (grade 3 and grade 4).

| n (%) | Untreated AML,<br>n=23 | Relapsed / Refractory AML,<br>n=18 |
| --- | --- | --- |
| Neutropenia | 23 (100) | 18 (100) |
| Febrile neutropenia | 19 (83) | 7 (39) |
| Anemia | 20 (87) | 15 (83) |
| Thrombocytopenia | 20 (87) | 17 (94) |
| Hepatic dysfunction | 0 (0) | 0 (0) |
| Renal Insufficiency | 1 (4) | 0 (0) |
| Nausea | 2 (9) | 0 (0) |
| Fatigue | 2 (9) | 1 (6) |
| Infection | 4 (33) | 3 (21) |
| Invasive fungal infection | 0 (0) | 2 (13) |

### Molecular characteristics

Seventeen patients had complex karyotypes (untreated: 10, R/R: 7). Mutations and gene fusions were investigated in 38 patients by targeted NGS (Figure 1C). NGS analyses were not performed in 3 patients from whom samples were unavailable. All patients harbored at least one mutation. *TET2* was the gene most commonly mutated (22 patients; untreated: 14, R/R: 8), followed by *KMT2D* in 14, *RUNX1* in 12, *TP53* in 9, *NCOR2* in 7, *TSC1* and *DNMT3A* in 5 each, and *KRAS*, *NRAS*, *ASXL1*, *PHF6*, *ARID1B*, *NPMI*, and *IDH1* in 4 each. An *FLT3*/ITD mutation was observed in 3. An *FLT3*/TKD mutation was observed in one R/R patient. No patient harbored both *FLT3*/ITD and *FLT3*/TKD mutations. Among *KMT2D* mutations, nucleotide substitutions predicted to yield the amino-acid substitutions G2493V (exon 31), C1424S (exon 15), G3354R (exon 34), P646R (exon 10), and E3910L (exon 39) were observed (Supplementary Figure 1A). None of these changes has been recorded in the gnomAD V2.1.1 ^28^ database or the National Center for Biotechnology Information (NCBI) database ^29^.

Of note is that one patient harbored the novel fusion gene *FHIT-TIRAP*. Whilst the *FHIT* FRA3B locus is fragile and has participated in many recorded gene fusions, *FHIT-TIRAP* is not among those in the gnomAD V2.1.1 database, the NCBI database, and the ChimerDB 4.0 database ^29^. Therefore, we consider this fusion gene novel.

### Survival

Median overall survival was 655 days in untreated patients. R/R patients had a median overall survival of 355 days (untreated *vs.* R/R, p= 0.1751) (Figure 1D).

We performed subgroup analyses using existing indexes, ELN risk stratification (favorable and intermediate *vs.* adverse), and presence of a complex karyotype. When comparing patients by ELN risk stratification, adverse risk had a median survival of 279 days, whereas median survival was 622 days for intermediate and favorable risk (p=0.0973). A complex karyotype significantly affects OS (p=0.0259) (Figure 1E). We further compared OS by use of G-CSF. Although OS did not differ significantly between those receiving G-CSF and those not, and although median survival was 425 days, G-CSF usage reduced the number of days that patients were affected by FN, thereby contributing to better outcomes. Comparisons of OS by subsequent HSCT found a statistically significant difference (p=0.0498). Median survival was 655 days for patients who underwent HSCT and 279 days for patients who did not undergo HSCT (Figure 1F).

In subgroup analyses of median survival in patients harboring mutations associated with adverse prognosis, individually and in combination. We evaluated *TP53, ASXL1+RUNX1*, *KRAS/NRAS, TET2, KMT2D, IDH1/2,* and *FLT3-*ITD*+*TKD. Presence of *TP53* alterations was an independent predictor of poor survival outcome. All 13 patients who harbored *TP53* alterations (*TP53* mutations, 9; loss of chromosome 17, 5) had complex karyotypes. The median survival of patients with *TP53* alterations was 146 days, significantly different (p*=*0.0008) from patients without *TP53* alterations (“wild-type”; median survival was 655 days) (Figure 1G). Other mutations did not significantly affect OS (Supplementary Figure 1B).

Multivariate analyses revealed that prior treatment (R/R status) and *TP53* alterations were associated with poorer outcome, whilst CR was associated with better outcome (Supplementary Figure 1C).

### TP53 mutation types and loss of function

Nine patients (22.0%; untreated: 7, R/R: 2) harbored *TP53* mutations (Figure 1C). Ten mutations (90.9%) were localized within the DNA binding domain and one (9.1%) within the hinge domain. *TP53* mutations were most commonly of missense type, accounting for 81.8%. The remaining 2 were truncating mutations. All *TP53* mutations detected resulted in loss of function of *TP53* (Figure 2A). “Single-hit” is defined as a single *TP53* mutation or loss of chromosome 17 (single *TP53* mutation: 6, loss of chromosome 17: 4) and “Double-hit” is defined as single *TP53* mutation with loss of chromosome 17, or double *TP53* mutations (single *TP53* mutation and loss of chromosome 17: 1, double *TP53* mutations: 2) in Figure 2B. Significant survival difference was also observed between single-hit and wild-type *TP53* (p = 0.0003). No significant difference was found between double-hit and wild-type *TP53*, possibly due to the small double-hit sample size.

**Figure 2.**
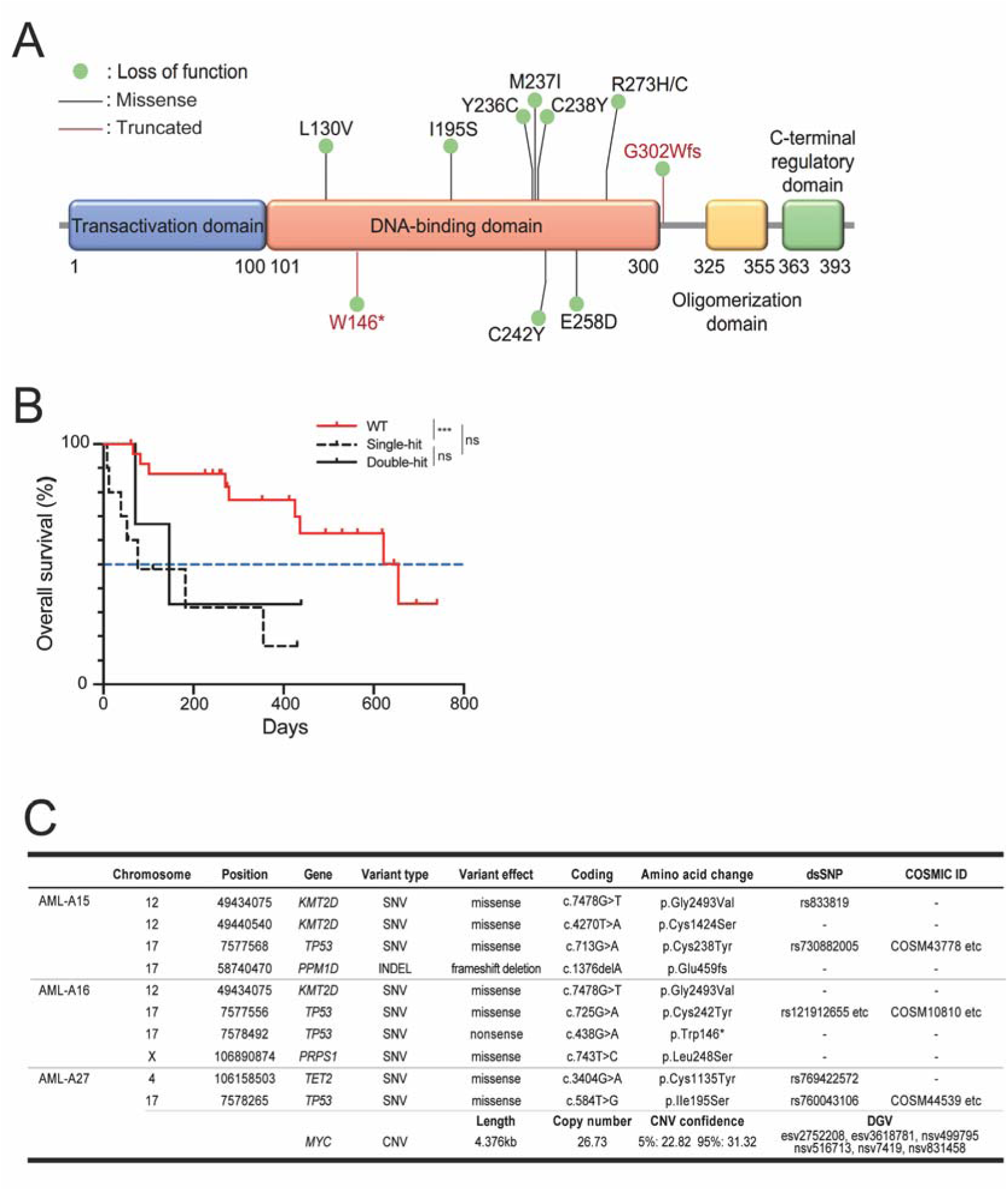
Prognostic significance of frequency of TP53 mutations. (A) Distribution of *TP53* mutations along the gene body in this study. (B) Overall survival of patients by frequency of *TP53* subgroups (WT; n=26, single-hit; n=10, double-hit; n=3). *** p<0.001. (C) Genetic mutations in three *TP53*-mutated patients (AML-15, 16, 27).

### Single cell RNA-sequence data processing and analysis

To investigate how venetoclax and azacitidine therapy affected tumor microenvironment of *TP53*-mutated AML, we analyzed gene expression by scRNA-seq in bone marrow samples obtained pre- and post-treatment from 3 untreated patients with *TP53*-mutated AML (AML-15, 16, 27) (Figure 2C). Post-treatment marrow was sampled 1 month after starting standard-dose venetoclax and azacitidine therapy. The status of AML-15 was assessed as CRi and that of AML-16 and −27 was assessed as CR.

Annotation of each cluster was determined by gene expression, heatmap analysis, and published descriptions of activity in peripheral blood or bone marrow ^26,27^ (Figure 3A). This yielded 12 individual clusters (Figure 3B), with Cluster 0: CD4 T cells, Cluster 1: CD8 T cells No. 1, Cluster 2: NK cells No.1/ CD8 T cells No.2, Cluster 3: NK cells No.2/CD8 T cells No.3, Cluster 4: AML cells No.1/Dendritic cells, Cluster 5: AML cells No.2, Cluster 6: AML cells No.3, Cluster 7: monocytes/AML cells No.4, Cluster 8: AML cells No.5, Cluster 9: AML cells No.6/CD8 T cells No.4, Cluster 10: B cells, and Cluster 11: plasma cells. Venetoclax and azacitidine therapy eradicated most immature cells (*CD34* and *KIT* population) (Figure 3C) a population clearly matching that of AML cells^30^. The expression of *WT1* also decreased post-treatment (Figure 3C).

**Figure 3.**
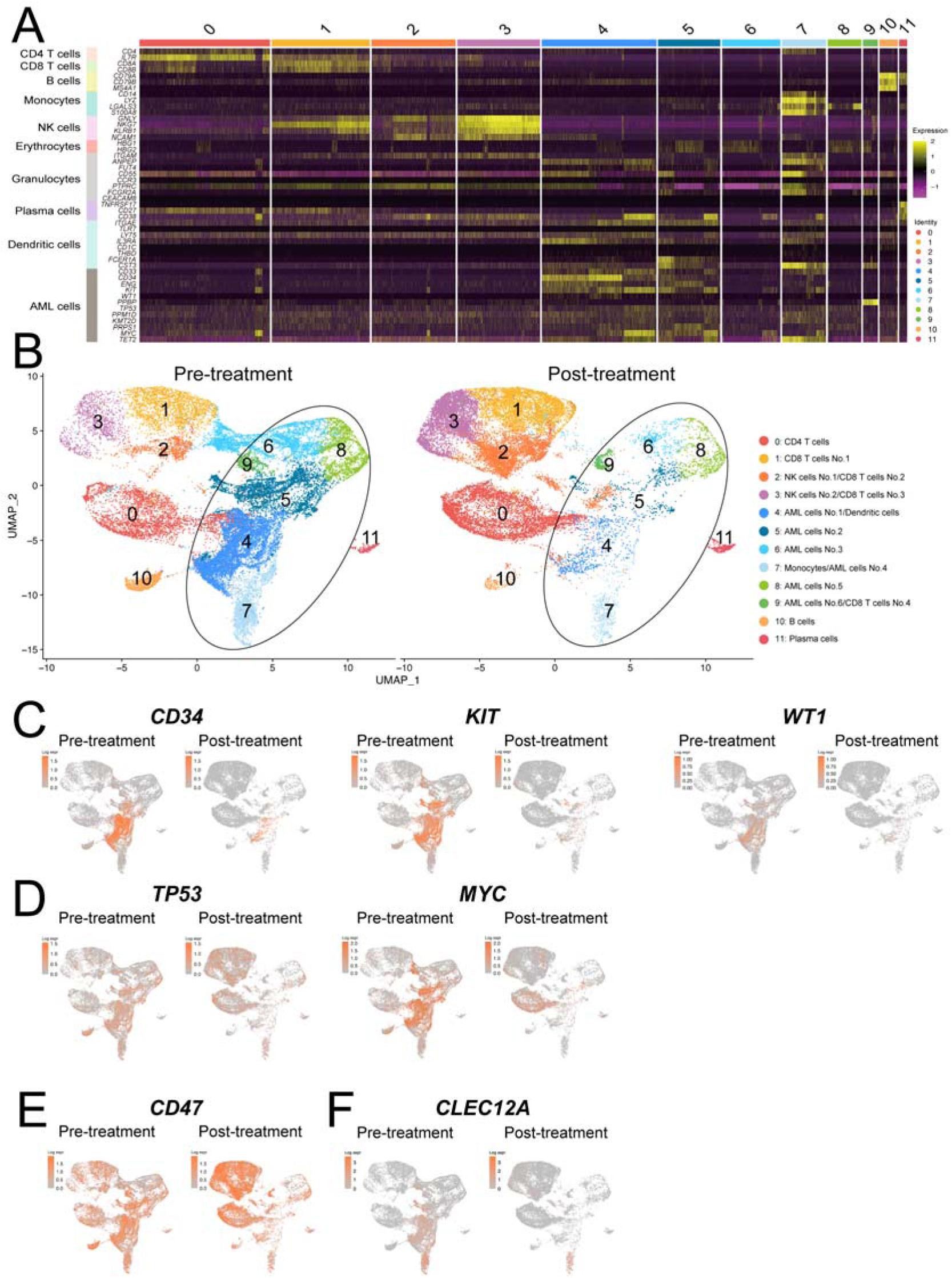
ScRNA-seq comparing *TP53* mutated patient bone marrow samples pre- and post-venetoclax and azacitidine treatment from 3 untreated patients with *TP53*-mutated AML (AML-15, 16, 27). (A) Heatmap of genotype groups composed of unique signature genes for each cluster. (B) Uniform Manifold Approximation and Projection (UMAP) of scRNA-seq data showing pre-treatment (left) and post-treatment (right). Cluster 0: CD4 T cells, Cluster 1: CD8 T cells No. 1, Cluster 2: NK cells No.1/ CD8 T cells No.2, Cluster 3: NK cells No.2/CD8 T cells No.3, Cluster 4: AML cells No.1/Dendritic cells, Cluster 5: AML cells No.2, Cluster 6: AML cells No.3, Cluster 7: monocytes/AML cells No.4, Cluster 8: AML cells No.5, Cluster 9: AML cells No.6/CD8 T cells No.4, Cluster 10: B cells, Cluster 11: plasma cells. (C-E) UMAP of individual genes associated with (C) (D)AML cells, and (E) *CD47* found in the patient studied (F) *CLEC12A* found in the patient studied.

AML-15 harbored mutations in *KMT2D, TP53*, *PPM1D*, AML-16 in *KMT2D, TP53*, *PRPS1*, and AML-A27 in *TET2*, *TP53*, *MYC* (Figure 2C). Expression levels of these genes were clearly decreased in clusters involving AML cells after venetoclax combination therapy (Figure 3D and Supplementary Figure 2A). *TP53* expression recovered within clusters 0, 1, 2, 3 and 10, which did not include AML cells (Figure 3D). CD47, a “don’t eat me” signal overexpressed on malignant cells (including AML cells) and a therapeutic target, was predominantly found pre-treatment and decreased in incidence post-treatment (Figure 3E). Furthermore, *CLEC12A* (CLL1) was expressed in cluster 7 pre-treatment and remained post-treatment (Figure 3F).

Numbers of cells expressing anti-apoptotic genes such as *BCL2*, *MCL1*, and *MDM2* decreased in clusters involving AML cells after venetoclax combination therapy (Figure 4A), although gene expression levels did not differ significantly (Supplementary Figure 2B). Cells expressing pro-apoptotic genes such as *BAX*, *BID*, and *ATM* also decreased in number due to elimination of most AML cells. On the other hand, non-AML cells recovered with high expression of these genes (Figure 4B).

**Figure 4.**
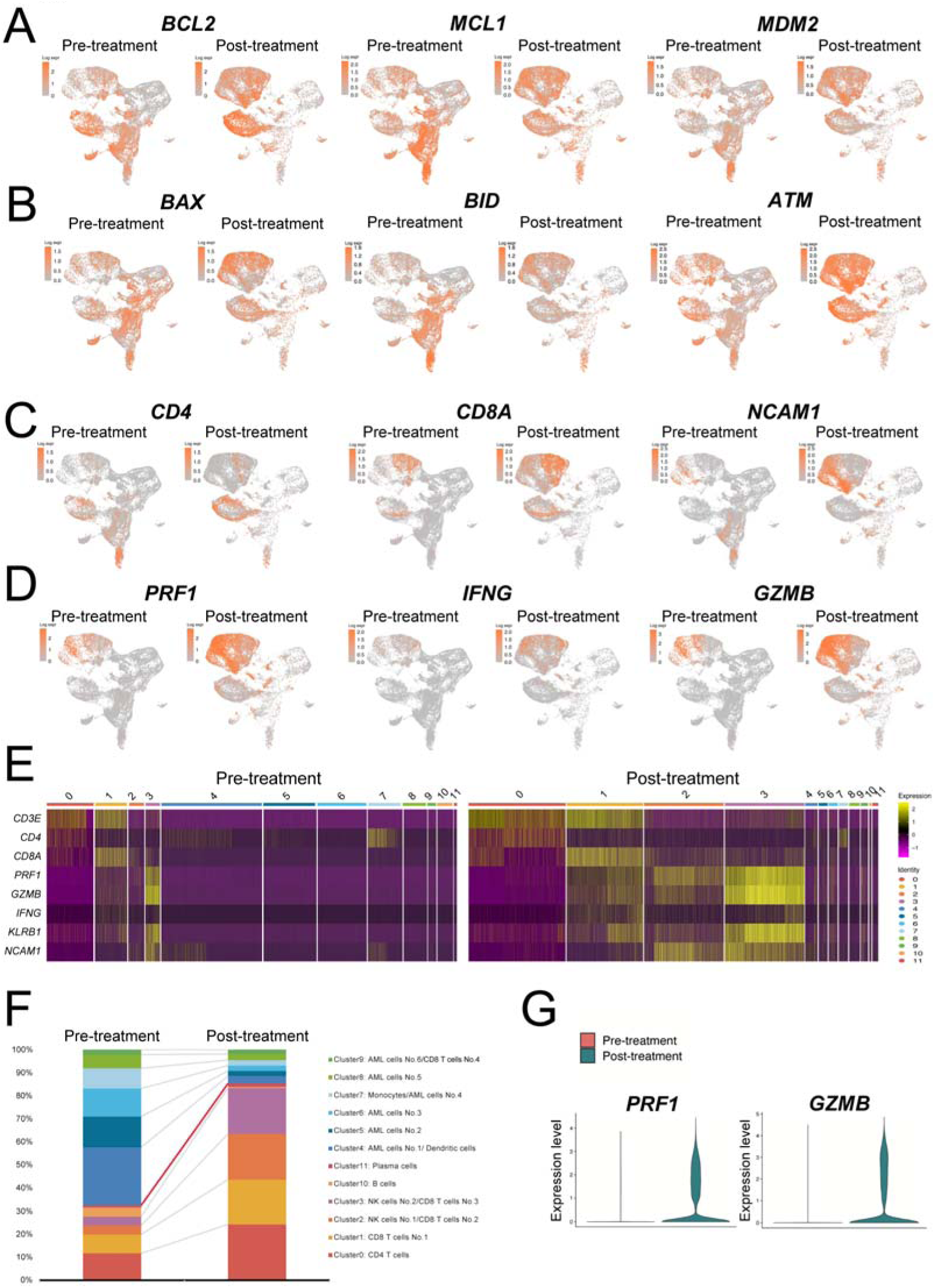
ScRNA-seq reveals suppression of anti-apoptotic potential and recovery of immune cells post-venetoclax and azacitidine treatment. (A) UMAP of genes associated with anti-apoptosis. (B) UMAP of genes associated with pro-apoptosis. (C-D) UMAP of genes associated with immune cells pre- and post-venetoclax and azacitidine treatment. (C) CD 4 T cells, CD 8 T cells, NK cells, (D) cytotoxicity. (E) Heatmap of genes associated with immune cells. (F) Ratio of clusters pre- and post-venetoclax and azacitidine treatment. (G) Violin plot of gene expression of *GZMB* and *PRF1* pre-and post-treatment.

Expression levels of genes associated with patterning the tumor microenvironment also were examined. CD8 T cells, CD4 T cells, and NK cells (*NCAM1, KLRB1*) were significantly increased both in number and in levels of gene expression post-treatment in all 3 patients (Figure 4C-G and Supplementary Figure 2C). Moreover, CD 8 T cells and NK cells, both present in increased numbers, expressed *PRF1*, *IFNG*, and *GZMB* (Figure 4E and 4G) at high levels, consistent with potential for cytotoxicity. These data indicate immune recovery and decreased numbers of cells expressing anti-apoptotic genes after venetoclax and azacitidine therapy.

## Discussion

Our observations in this study mirror the results of the VIALE studies ^11,12^ and of real-world reports ^13–20,31–35^. With R/R patients in particular, an ORR of 61.1% was an encouraging result. In most of our patients venetoclax doses were not decreased when antifungal prophylaxis was administered. The incidence of invasive fungal infections (IFI) in AML patients during venetoclax combination therapies is reported as 5 – 13% ^21,36,37^. In our study, only 2 patients (4.88%) received antifungal prophylaxis, but only 2 patients, including the patient given voriconazole, had IFI. Our experience is consonant with that of others whose AML patients did not differ significantly in IFI incidence due to antifungal prophylaxis ^21,36^. We accordingly suggest that antifungal prophylaxis is not necessarily required for all patients before venetoclax combination therapy is begun, bearing in mind that to deploy antifungal treatment when it is needed is very important. OS may have been improved by proactive use of G-CSF in managing cytopenia. Episodes of grade ≥3 neutropenia and FN generally were shorter in patients given post-remission G-CSF than in patients who were not, although no statistically significant difference was demonstrated (Figure 1F). Optimal treatment of FN together with proactive use of G-CSF for cytopenia management may have contributed to better OS.

Mutations have been reported to affect prognosis of AML patients receiving this treatment ^23^: More specifically, mutation in *NPM1* is associated with better outcome ^19,22,32,38^, whereas mutations in *RUNX1, ASXL1, TP53*, the RAS pathway (*KRAS/NRAS*), and *FLT3* (*FLT3-* ITD*/*TKD) are associated with poor outcome ^17,19,22,39–41^. We detected at least one mutation in all patients, perhaps because use of both a multigene panel and an *FLT3*/ITD mutation detection kit let us search broadly.

Among the mutations assessed, only *TP53* alterations predicted poor outcome (Figure 1G). Median survival for patients with mutated *TP53* and complex karyotype who undergo HSCT was 4.8 months in one study, with 38% of patients dying before post-transplantation day 100 and >80% dying within 2 years after transplantation ^41^. With survival prognoses of 4.8 months for HSCT and 4.87 months for venetoclax combination therapy, which can be administered on an outpatient basis after the 2^nd^ cycle, the latter may have a significant advantage in treatment choice (improved patient quality of life).

In AML, macrophage-induced antibody-dependent cellular phagocytosis has been demonstrated *in vitro* on dual treatment with CD47-targeting antibodies and azacitidine ^42,43^. The combination of venetoclax and therapeutic antibodies increases *in vitro* phagocytosis in an apoptosis-independent manner ^44,45^. Triplet therapy (venetoclax, azacitidine, and anti-CD47 antibody) thus could improve prognosis in AML with *TP53* mutation. In our study, expression of *CD47* in *TP53*-mutated AML cells fell after only a single course of venetoclax and azacitidine. However, a small population of *CD47*-expressing AML cells remained. CD47-directed therapy might eradicate minimal residual disease. Furthermore, *CLEC12A* (CLL1)-expressing AML cells remained post-treatment in cluster 7. CLL1 is expressed on AML cells, but not on normal hematopoietic cells, and has been reported to be associated with quick relapse ^46^. CLL1-directed chimeric antigen receptor T cell therapy has shown promise in R/R AML ^47^, and may eradicate minimal residual disease in *TP53*-mutated AML cells after venetoclax combination therapy as well.

In conclusion, our study demonstrated encouraging responses to venetoclax combination treatment in untreated and R/R AML patients. This notwithstanding, the presence of *TP53* alterations was clearly an independent predictor of poor outcome. Further research is needed to improve OS in unfit AML patients, especially those with *TP53* alterations. We found in patients that cytotoxic CD8 T cells, CD4 T cells, and NK cells clearly increased both in number and in levels of gene expression associated with cytotoxicity post-treatment, resulting in immune recovery. CD47 and CLL1 was predominantly found on AML cells, which persisted post-treatment, permitting the inference that additional CD47- and CLL1-directed therapy may be a promising therapeutic option in *TP53*-mutated AML.

## Data Availability

All data produced in the present study are available upon reasonable request to the authors.

## Acknowledgements

We thank A.S. Knisely for critical reading of the manuscript.

## Authorship contributions

K.Y. performed scRNA-seq and wrote the manuscript. J.A. planned the study, supplied clinical data, and provided scientific discussions. Y.F. performed NGS and scRNA-seq, analyzed data, and wrote the manuscript. M.I. performed NGS and scRNA-seq and analyzed data. S.K. helped with performing NGS and scRNA-seq. K.T. analyzed scRNA-seq data and provided scientific discussions. Y.A. helped with writing the manuscript. Y.E., Y.T., H.Y., T.T., Y.H., and M.S. provided scientific discussions. M.N. conducted Kaplan-Meier analyses. M.A. planned and directed the study, analyzed data, and wrote the manuscript.

---

Supplementary Figure 1. Subgroup analyses of overall survival presence of mutations and multivariate analyses. (A) *KMT2D* mutations not recorded in gnomAD and NCBI databases. (B) Subgroup analyses of overall survival presence of mutations associated with adverse prognosis, individually and in combination. (C) Multivariate analyses affecting OS.

Supplementary Figure 2. AML cells post-treatment show decrease in gene expression and in AML cell number (A) UMAP of individual genes associated with AML cells. (B) Violin plot of gene expression of *BCL2, MCL1, MDM2,* pre-and post-treatment. (C) The number of cells in each cluster for three patients.

