## Supplementary Figures for "Immune recovery in tumor microenvironment of *TP53*-mutated AML following venetoclax combination therapy"

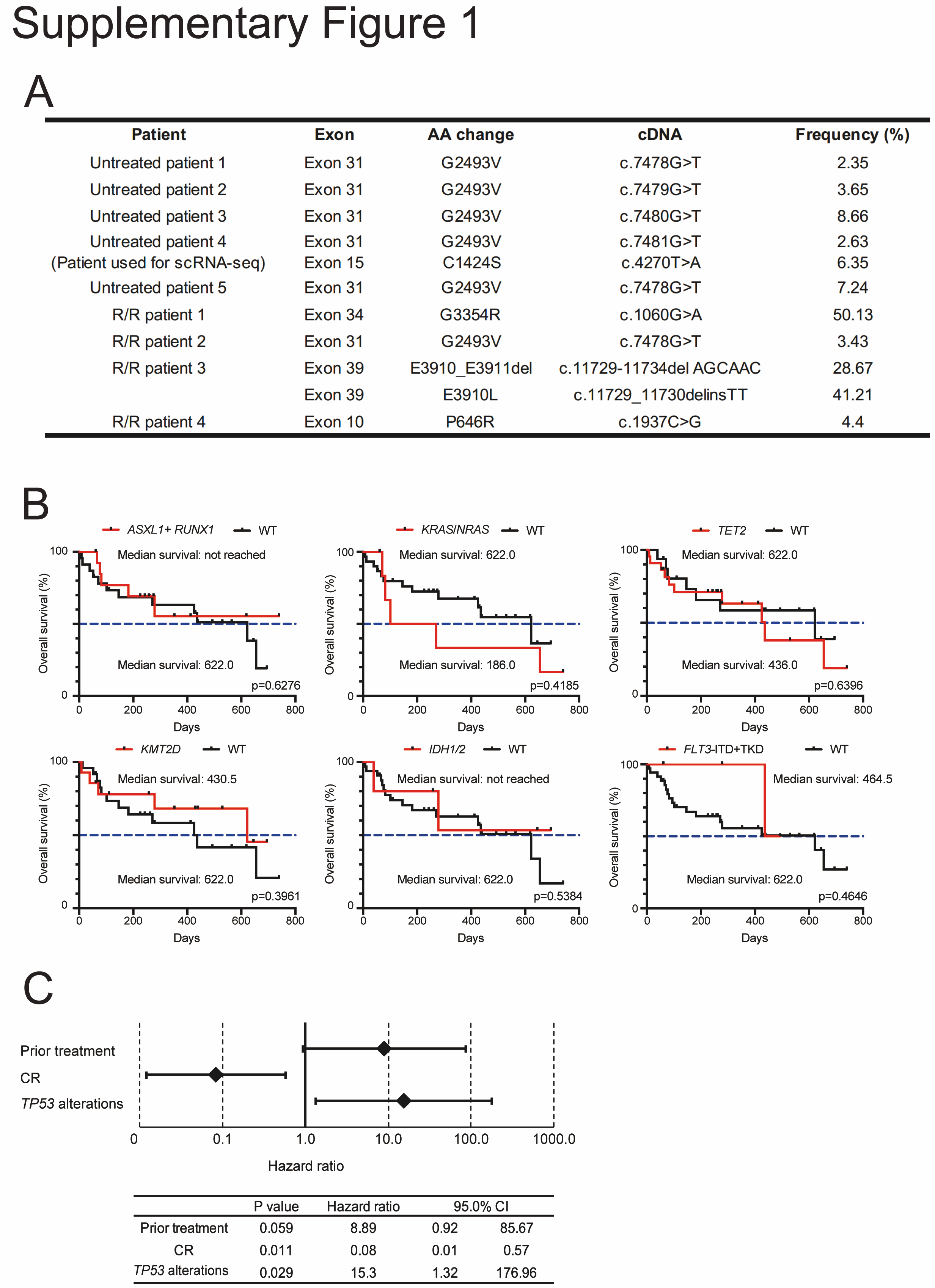

Supplementary Figure 1. Subgroup analyses of overall survival presence of mutations and multivariate analyses. (A) *KMT2D* mutations not recorded in gnomAD and NCBI databases. (B) Subgroup analyses of overall survival presence of mutations associated with adverse prognosis, individually and in combination. (C) Multivariate analyses affecting OS.

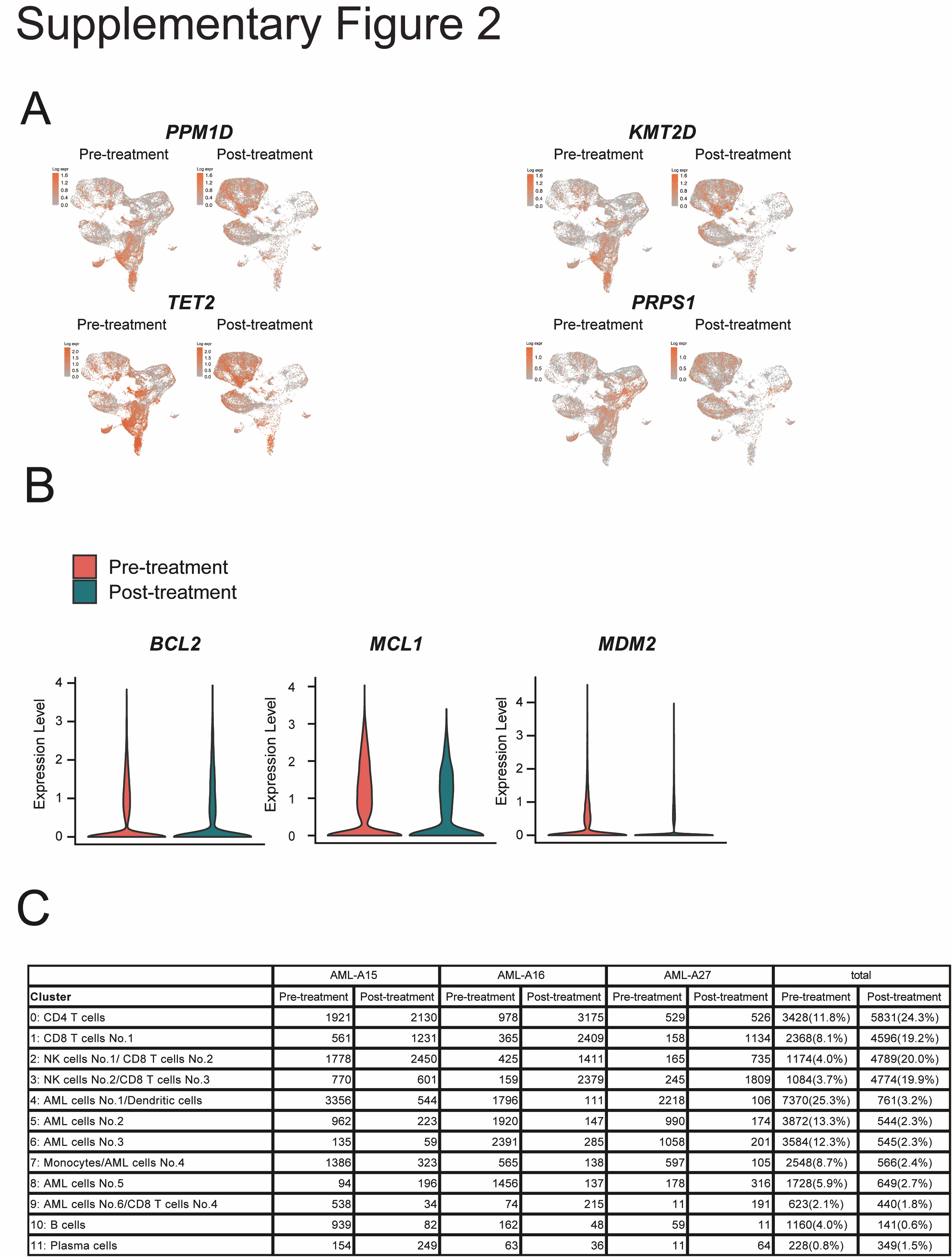

Supplementary Figure 2. AML cells post-treatment show decrease in gene expression and in AML cell number (A) UMAP of individual genes associated with AML cells. (B) Violin plot of gene expression of *BCL2, MCL1, MDM2,* pre-and post- treatment. (C) The number of cells in each cluster for three patients.
